# Hospitalization forecast to inform COVID-19 pandemic planning and resource allocation using mathematical models

**DOI:** 10.1101/2022.11.03.22281898

**Authors:** Philip Erick Wikman-Jorgensen, Angel Ruiz, Vicente Giner-Galvañ, Jara Llenas-García, José Miguel Seguí-Ripoll, Jose María Salinas Serrano, Emilio Borrajo, José María Ibarra Sánchez, José Pedro García-Sabater, Juan A Marín-García

## Abstract

**Background:** The COVID-19 pandemic has put tremendous pressure on hospital resources around the world. Forecasting demand for healthcare services is important generally, but crucial in epidemic contexts, both to facilitate resource planning and to inform situational awareness. There is abundant research on methods for predicting the spread of COVID-19 and even the arrival of COVID-19 patients to hospitals emergency departments. This study builds on that work to propose a hybrid tool, combining a stochastic Markov model and a discrete event simulation model to dynamically predict hospital admissions and total daily occupancy of hospital and ICU beds.

**Methods:** The model was developed and validated at San Juan de Alicante University Hospital from 10 July 2020 to 10 January 2022 and externally validated at Hospital Vega Baja. An admissions generator was developed using a stochastic Markov model that feeds a discrete event simulation model in R. Positive microbiological SARS-COV-2 results from the health department’s catchment population were stratified by patient age to calculate the probabilities of hospital admission. Admitted patients follow distinct pathways through the hospital, which are simulated by the discrete event simulation model, allowing administrators to estimate the bed occupancy for the next week. The median absolute difference (MAD) between predicted and actual demand was used as a model performance measure.

**Results:** With respect to the San Juan hospital data, the admissions generator yielded a MAD of 6 admissions/week (interquartile range [IQR] 2-11). The MAD between the tool’s predictions and actual bed occupancy was 20 beds/day (IQR 5-43), or 5% of the hospital beds. The MAD between the intensive care unit (ICU)’s predicted and actual occupancy was 4 beds/day (IQR 2-7), or 25% of the beds. When the model was further evaluated with data from Hospital Vega Baja, the admissions generator showed a MAD of 2.42 admissions/week (IQR 1.02-7.41). The MAD between the tools’ predictions and the actual bed occupancy was 18 beds/day (IQR 19.57-38.89), or 5.1% of the hospital beds. For ICU beds, the MAD was 3 beds/day (IQR 1-5), or 21.4% of the ICU beds.

**Conclusion:** Predictions of hospital admissions, ward beds, and ICU occupancy for COVID-19 patients were very useful to hospital managers, allowing early planning of hospital resource allocation.

## Background

In 2020, the World Health Organization (WHO) declared COVID-19 a public health emergency of international concern.[1] Since then, it has turned out to be the most important pandemic in a century. From an economic standpoint, numerous studies report staggering impacts. In Spain, a study in six long-term care facilities estimated that direct medical costs amounted to EUR 276,281 per month during the epidemic.[2] In another study in Madrid, the cost of medical treatments reached EUR 440,000 per 1000 hospitalized patients. Nevertheless, the 10.8% drop in the gross domestic product (GDP) is the most demonstrative figure of the economic fallout from the pandemic.[3] In the USA, total direct medical costs throughout the pandemic have been estimated to range from USD 163.4 to USD 654 billion, depending on the total amount of people finally infected.[4] Similar repercussions have been reported in Turkey, Switzerland, and China.[5–7]

The pandemic has also had unprecedented impacts on global health systems, with administrators and doctors struggling to allocate sufficient resources to meet the demand for services.[8] During the first wave in early 2020, hospital capacity was dramatically overwhelmed, not only in Spain but worldwide.[9] [10] To cope with the deficit of resources, planning and preparation measures to scale up hospital resources were implemented. Surgical theatres were transformed into intensive care units (ICU), while meeting rooms, libraries, and rehabilitation gyms were all refitted to serve as conventional wards. The stressful working conditions brought on by the pandemic also provoked substantial mental health problems among health care workers.[11]

The public health crisis put enormous pressure on administrators, who had to search for more medical resources and supplies, including conventional hospital beds, ICU beds, ventilators, protective personal equipment, and health care staff. Over the successive waves, administrators have also been challenged with decisions on when and how to scale down the deployment of organizational resources in order to return to normal activity levels as soon as possible.

Mathematical models have been widely used to inform public health policy during the pandemic.[12]Various epidemiological models have been developed;[13–15] however, these models address the dynamics of the pandemic at a regional or national level, so they are of limited use at the local or center level, where daily activity is organized. Models to predict the need for specific hospital resources have also been developed in the USA, Chile, and Europe.[16,17] [18][19] Nevertheless, the infrastructure and technical skills needed can be prohibitive.

In an effort to help support hospital administrators in resource planning for real short-term future pandemic-related needs, our group created a predictive tool based on local data. The aim of this study is to assess the predictive capacity of this local tool based on the real evolution of the pandemic in two similar Spanish hospitals.

## Methods

### Setting

This study took place in two hospitals in Alicante, Spain. The Hospital Universitario San Juan de Alicante serves 223,962 inhabitants and has 386 hospital beds, of which 16 are ICU beds. The Hospital Vega Baja has a catchment population of 167,149 inhabitants and has 350 total beds and 14 ICU beds.

### Study population

We aimed to predict new hospital admissions and the need for hospital beds and ICU beds for patients infected with COVID-19. The study included all people in the hospital catchment area with a positive microbiological COVID-19 test from 10 July 2020 to 10 January 2022. No exclusion criteria were applied.

### Predictive tool

Figure 1 shows the complete patient pathway. Briefly, the admissions generator determines whether a COVID-19 patient that tests positive is likely to need hospitalization. Once admitted, they may or may not need ICU admission and then are finally discharged (dead or alive).

**Figure 1.**
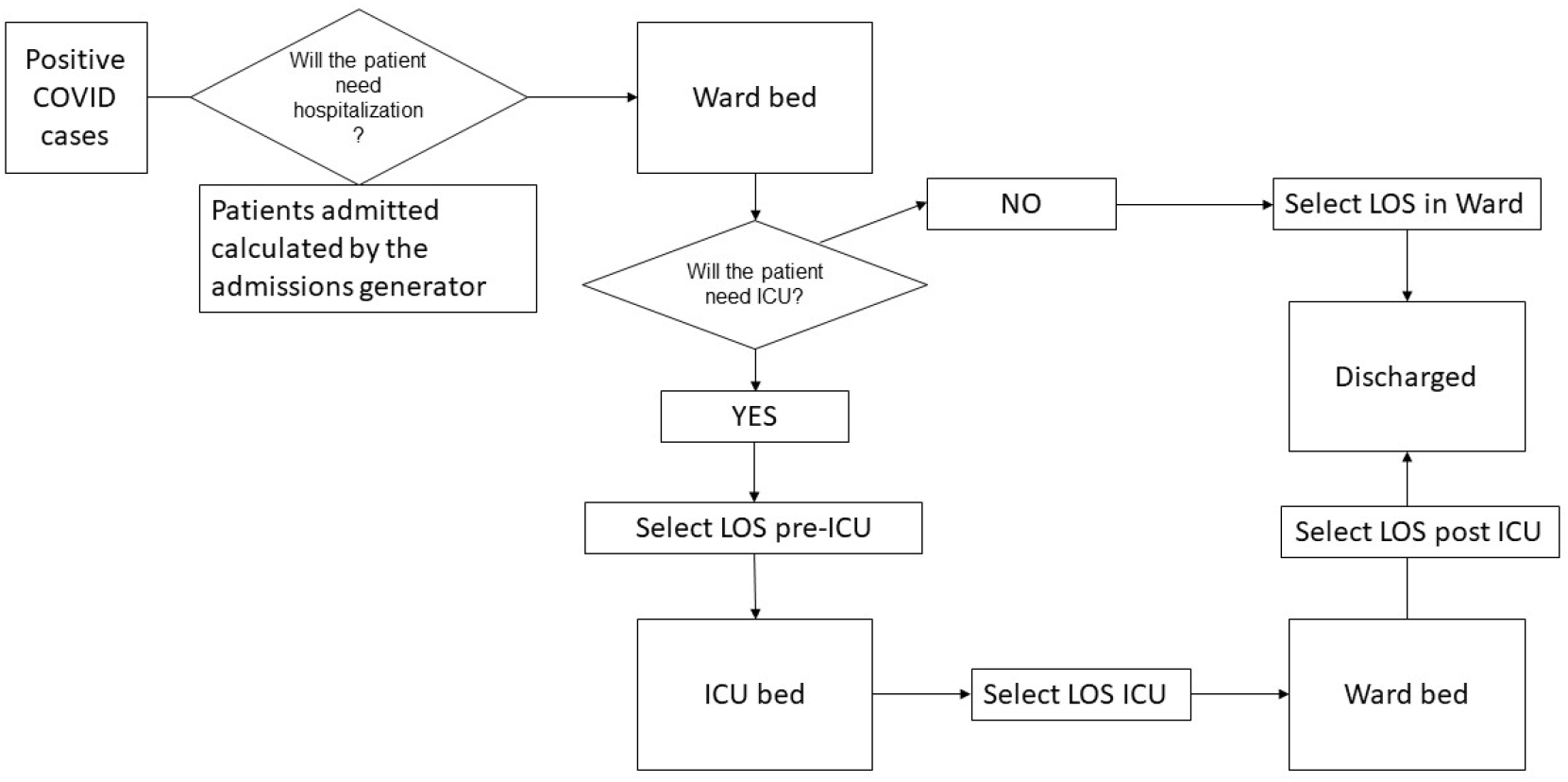
Schematic representation of the forecast tool. ICU: intensive care unit; LOS: length of stay.

### Admissions generator

We developed a two-step Markov model to predict the likelihood of hospitalization in the general population with a microbiologically confirmed COVID test (either PCR or antigen test). To this end, the age-adjusted probabilities of hospitalization as reported by Verity et al.[20] were used. The probability of admission was multiplied by the number of positive tests within each age group. Infected patients who need admission are usually hospitalized around the seventh day of symptoms, so assuming that the test is done on the first day of symptoms onset and that all the patients tested in the area served by the hospital are processed at its Microbiology Department, it is possible to estimate the number of patients that need hospitalization one week in advance. Due to variability in the natural course of the disease in each patient as well as a weekend effect in the testing chain, the total number of hospitalizations was calculated on a weekly rather than daily basis. This allowed us to predict the expected number of total hospitalizations from one week to the next. As of 15 December 2021, the probability of hospitalization fell by 70%, corresponding with the risk reduction due to vaccination; this figure was adjusted for a vaccination coverage of 80%.[21]

### Hospitalization pathway

To model the length of stay on a conventional ward, the data for the length of stay at the Hospital Universitario San Juan de Alicante was used. The length of stay was fitted to a Weibull distribution using Neld-Meld optimization method as implemented in the R stats package. To model the length of stay in the ICU, the same procedure was undertaken.

A discrete event simulation model was programmed in R using the simmer package.[22,23]. The model was fed by the generator. Length of hospital and ICU stay were modelled using the cited distributions. The code is available at https://github.com/pwjpwj/COVID upon reasonable request to the corresponding author.

To assess the performance of the predictions, the absolute difference between predicted and observed occupancies were measured and reported as the median absolute difference (MAD) with their interquartile range (IQR). Real data on admissions, hospital bed occupancy, and ICU bed occupancy were obtained from the hospital system’s electronic medical records.

From 10 July 2020 to 10 January 2022, the model was used to guide hospital policy and informed by a continuous and simultaneous validation of results.

To evaluate whether other hospitals could make use of the tools, a second test was conducted at the Hospital Vega Baja during the same period.

## Results

The admissions generator was programmed in R and validated through visual inspection of the predictions and comparison to real admissions, as shown in Figure 2. As a metric of accuracy, the MAD between prediction and reality was 6 admissions/week (IQR 2-11).

**Figure 2.**
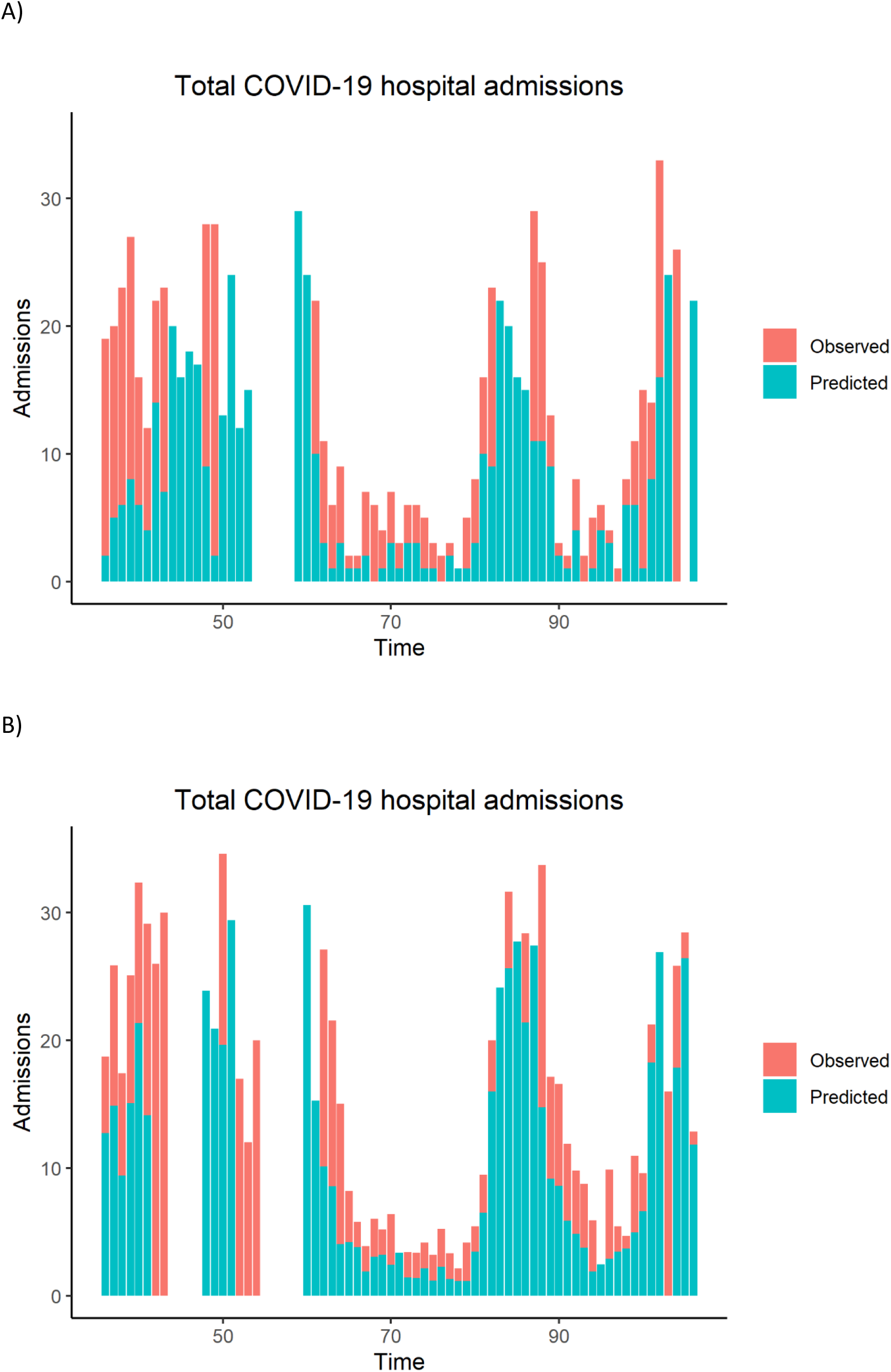
Comparison of predicted weekly number of admissions and actual admissions over the observation period in the Hospital Universitario de San Juan (A) and the Hospital Vega Baja (B). Time is represented as weeks since 1 September 2020. Blank columns correspond to periods without admissions.

The hospitalization pathway was programmed in R. Length of stay in a conventional ward and in the ICU was modelled using a Weibull distribution (Table 1) and validated by visual inspection (Figure 3). As a metric of accuracy, the MAD between prediction and reality was 20 beds/day (IQR 5-43), corresponding to 5% of total hospital beds. Real and predicted ICU bed occupancy was also evaluated (Figure 4), yielding a MAD of 4 beds (IQR 2-7), or 25% of ICU capacity.

**Table 1.** Model probabilities and data

|  | **Estimate** | **Distribution** | **Reference** |
| --- | --- | --- | --- |
| Age-stratified probability of hospitalization |  | - | Verity et al.[17] |
| 0-9 years | 0 |  |  |
| 10-19 years | 0.000408 |  |  |
| 20-29 years | 0.0104 |  |  |
| 30-39 years | 0.0343 |  |  |
| 40-49 years | 0.0425 |  |  |
| 50-59 years | 0.0816 |  |  |
| 60-69 years | 0.118 |  |  |
| 70-79 years | 0.166 |  |  |
| >80 years | 0.184 |  |  |
| Probability of ICU admission | 0.15 | - | Berenguer et al. [23] |
| Length of hospital stay | Shape=1.28<br>Scale=10.61 | Weibull | Own measurements |
| Time in normal ward before ICU | Shape=1.52<br>Scale=5.4 | Weibull | Own measurements |
| Time in normal ward after ICU | Shape=1.39<br>Scale=17.05 | Weibull | Own measurements |
| Time in ICU | Shape=1.06<br>Scale=19.24 | Weibull | Own measurements |

**Figure 3.**
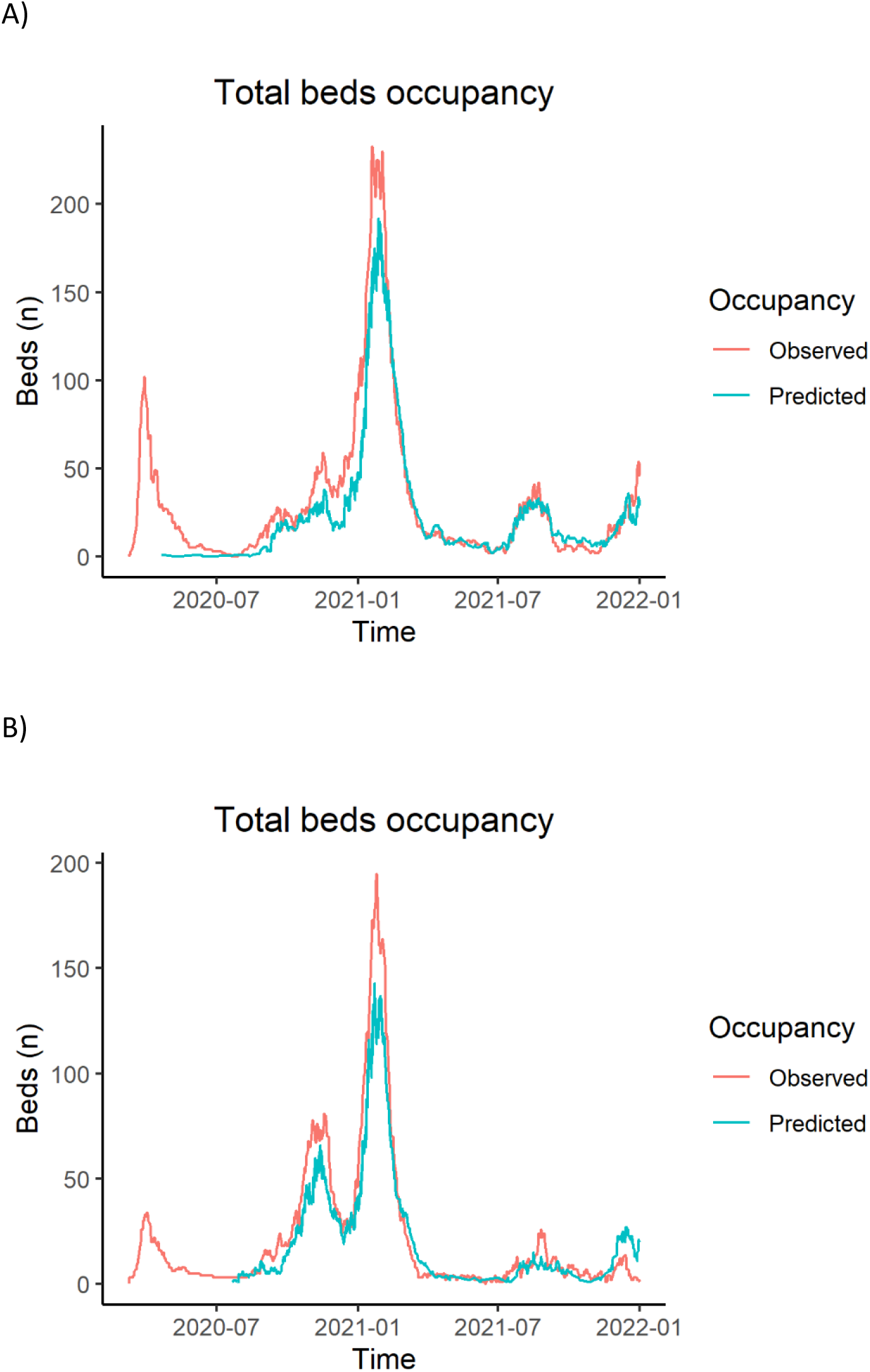
Comparison between predicted hospital bed occupancy and actual occupancy. A) Hospital Universitario San Juan de Alicante, B) Hospital Vega Baja.

**Figure 4.**
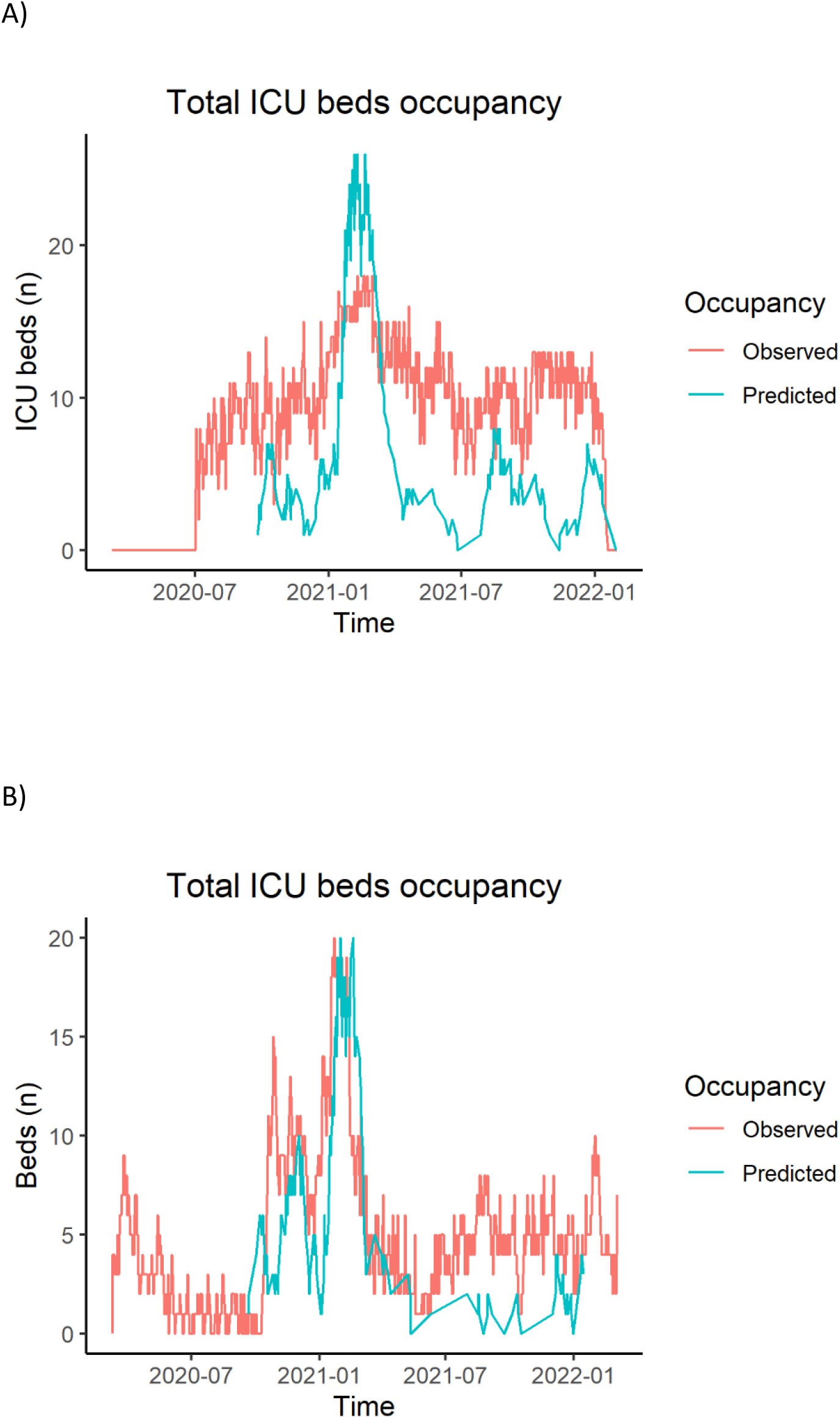
Comparison between predicted ICU bed occupancy and actual occupancy. A) Hospital Universitario San Juan de Alicante, B) Hospital Vega Baja.

A second assessment of the model was performed for Hospital Vega Baja. In this case, the admissions generator showed a MAD of 2.42 admissions/week (IQR 1.02-7.41). The hospitalization pathway simulator showed a MAD of 18 total hospital beds/day (IQR 19.57-38.89), 5.1% of capacity, and 3 ICU beds/day (IQR 1-5), 21.4% of capacity.

## Discussion

Using a prospective model based on the positive COVID-19 test results in the hospital’s catchment population, we were able to predict, one week in advance: 1) new hospitalizations, 2) the expected total number of hospital beds needed to attend these patients, and 3) the number of ICU beds needed. This information was used to plan and prepare the local resources needed in advance. The validity of the model was confirmed at a second hospital.

Previous models described in the literature [16,17] have achieved better accuracy for predicting admissions, but they are considerably more complicated to implement than our approach.[18,24] The very simple method of multiplying the number of COVID-19 positive tests by the age-stratified probability of hospitalization yielded useful estimates of future admissions at one week.

The use of a discrete event simulation model to predict not only new admissions, but also the total beds needed (through a complex dynamic composite calculation of new admissions, patients already on the ward, and patients discharged) is the most original feature of our work. Previous reports have used this approach [25], but validation was not reported as in our model. Recently, a discrete event simulation model similar to our approach has been described.[26] However, it modeled two entire autonomous regions in Spain and did not focus on a specific hospital. That study used a population growth Gompertz model to simulate patient admissions. This would preclude the need for patient-level microbiology data, which can be challenging to obtain in some settings.

The accuracy of our model is not perfect, and, for admissions, it does not perform as well as other models.[17] Moreover, the precision for ICU beds is worse than that for the total number of beds. However, our administrators considered it useful for improving planning and preparation.

One strength of the tool is that it is data-driven, making it less prone to errors. Also, the tool was developed at one center and validated at another, showing that the results are quite robust and work well in more than one setting. This external validation, together with its simplicity, makes it easy to scale up and apply elsewhere, with few infrastructure needs. The admissions generator can be implemented on an open-source spreadsheet. The discrete event simulation model tool, although somewhat more elaborate, can also be implemented with free software.

From a practical point of view, our model supported hospital resource planning on a local scale during successive COVID-19 waves. Additionally, it might also aid decision-making around when and how to reactivate elective surgical procedures suspended during peak epidemic periods.

In conclusion, the tool presented here enables the prediction of regular and ICU hospital admissions one week in advance for COVID-19 patients at a hospital level. The tool has been useful for improving planning and resource allocation at our centers and has been a source of reassurance for hospital staff in the face of the psychological stress of the unknown. Future developments will be needed to adjust for vaccine efficacy and coverage and emergence of new variants as well as to refine the tool’s precision and develop an admissions generator that does not rely on laboratory test results.

## Data Availability

Data is accesible at https://github.com/pwjpwj/COVID

